# Rural–urban disparities and associated factors of SARS-CoV-2 infection in Zambia: A convergent mixed-methods study using the Proximate Determinant Framework

**DOI:** 10.64898/2026.08.25.26361355

**Authors:** Enock Wantakisha, Stanley Nyirenda, Mony Narayani

**Affiliations:** Department of Public Health and Epidemiology, Chreso University, Lusaka, Zambia; Department of Immunology and Pathology, Central Veterinary Research Institute, Lusaka, Zambia; School of Public Health, Texila American University, Lusaka, Zambia

## Abstract

**Background:** Rural-urban disparities in SARS-CoV-2 infection epidemiology remain poorly quantified and understood in Zambia despite differences in healthcare access, services and preventive interventions. This study examined the geographical distribution and associated factors of SARS-CoV-2 cases across selected rural and urban districts of Zambia.

**Methods:** A convergent mixed-methods study comprised of quantitative survey and qualitative interviews was conducted in; Ndola (Urban), Kafue (Peri-urban) and Lufwanyama (Rural). The proximate determinant framework guided variable selection and interpretation. Quantitative combined (Hospital-surveillance data with community survey), while qualitative included In-depth interviews. Participants were sampled using multistage sampling technique. Quantitative data were analysed using STATA version 17, while qualitative data were analysed thematically. Findings were integrated through triangulation.

**Results:** A total of 528 participants were included, with a median age 31 years (15–71). Overall SARS-CoV-2 positivity was 12.6%, varying across rural (16.5%), peri-urban (14.9%), and urban (9.9%) settings, though residence was not associated with infection (P<0.132). Participants aged ≥49 years had significantly higher odds of infection (aOR=8.78; 95% CI:1.15–66.99), whereas secondary education (aOR=0.37; 95% CI:0.16–0.86) and hospital-based testing (aOR=0.37; 95% CI:0.15–0.92) were associated with lower odds of infection. Vaccine uptake was highest in urban areas but was not independently associated with infection. Qualitative findings revealed marked rural–urban differences in perceived susceptibility, testing access, vaccine decision-making, and adherence to preventive measures, explaining several quantitative observations.

**Conclusion:** SARS-CoV-2 infection across rural and urban settings in Zambia was influenced by demographic, behavioral, and health-system factors rather than geographic residence alone. These findings highlight the need for context-specific prevention strategies, equitable access to testing, strengthened community surveillance, and targeted risk communication to improve preparedness and response for future respiratory disease outbreaks.

## Introduction

Infectious global health diseases have continued to be a challenge in control despite many strategies implemented (1). Severe Acute Respiratory Syndrome Coronavirus 2 (SARS-COV-2) infection, a contagious viral infection accounted for over 1.2 million deaths globally between 2019-2021 (1–4). The pandemic exposed weak health systems, need for strengthened emergency preparedness and response (4–6). Post-pandemic isolated incidences are still being reported worldwide (6, 7).

Geographical incidences continue being recorded among high-risk populations though actively monitored (4, 8–10). Despite these efforts, global threat still exists with new variants reported in many countries (11–13). The impact of this genetic diversity in public health necessitates targeted-surveillance strategies that systematically sample and characterize predominant strains among populations whilst concomitantly monitoring proximate indicators and other factors that may be associated with the infection.

Few studies conducted in sub-Saharan Africa have compared epidemiological diversities across rural-urban settings using integrated mixed-method approaches. In addition, majority lack integration of epidemiological surveillance data and community survey to explain rural-urban disparities using the proximate determinant framework. This framework guided variable selection, analysis and interpretation. Existing Zambian studies have described SARS-CoV-2 epidemiology (14) and COVID-19 behaviours (9), but few have directly compared rural, peri-urban and urban contexts while integrating surveillance data, community-level evidence and qualitative explanations within a theoretically informed framework (2, 27). In Zambian context, rural-urban structural, diagnostic, social health barriers disparities (6, 7), alongside greater reliance on traditional, cultural and community health beliefs exists (8).

Focus for this study was to examine the rural-urban distribution, contextual and health system factors to SARS-CoV-2 infection by integrating community-based surveys and Hospital-based surveillance data. Specifically, the study aimed to describe geographic distribution of SARS-CoV-2 infection post-pandemic phase; Identify demographic, behavioural and health-system factors associated with the infection; and to integrate quantitive and qualitative findings to explain the rural-urban disparities in infection patterns.

## Methods

### Study design and setting

A convergent mixed-methods design integrating quantitative and qualitative approaches concurrently was employed (15, 16). Quantitative approach combined community survey with retrospective hospital-based surveillance data, while qualitative included In-Depth Interviews (IDIs). Both were analysed separately and later triangulated to identify convergence, complementary, and divergence findings.

Study was conducted in three districts of Zambia. Sites purposively selected to represent different urban-rural settlement and contextual variations. Ndola (Urban) district recorded higher numbers of cases in all epidemic waves comparatively while Kafue (Peri-urban) predominantly industrial setting experienced disease clusters in 2022. Lufwanyama (Rural) recorded low SARS-CoV-2 infection incidences comparatively. All eligible community members in sites and hospital-based data at Ndola Central Hospital-Level 3 (Ndola), Lufwanyama district Hospital-Level 1 (Lufwanyama) and Kafue General Hospital-Level 2 (Kafue) were selected and followed.

### Study population and selection criteria

Population included individuals above 15 years residing in study settings. Hospital-surveillance data included laboratory confirmed SARS-CoV-2 test results among other variables while community survey never collected SARS-CoV-2 infection test status. In-depth interviews included key informants; survivors of SARS-CoV-2 infection, health care providers; and health administrators. Cases (active) found during data collection were included as hospital-surveillance data.

### Proximate-determinant framework and SARS-CoV-2 infection dynamics

The framework is widely used in population health, epidemiology, and demographic research, particularly to explain how social, environmental, behavioural, and biological factors operate through intermediate mechanisms to influence health outcomes. The framework, Fig 1 guided variable selection, analysis and interpretation. According to the framework, underlying social, demographic, and structural factors do not directly cause infection but influence behavioural and biological pathways that determine disease occurrences. In this study, demographic characteristics (age, sex, education, marital status), geographical residence (urban, peri-urban and rural), and health-system factors were considered underlying determinants. These influenced proximate behavioural determinants including knowledge, perceived susceptibility, vaccination uptake, health-seeking behavior, access to testing, and adherence to preventive measures. These behavioural determinants subsequently affected biological exposure to SARS-CoV-2 infection, ultimately resulting in infection. Primary outcome of interest was SARS-CoV-2 infection status (Binary) defined as laboratory evidence of SARS-CoV-2 infection (PCR or serology).

**Fig 1:**
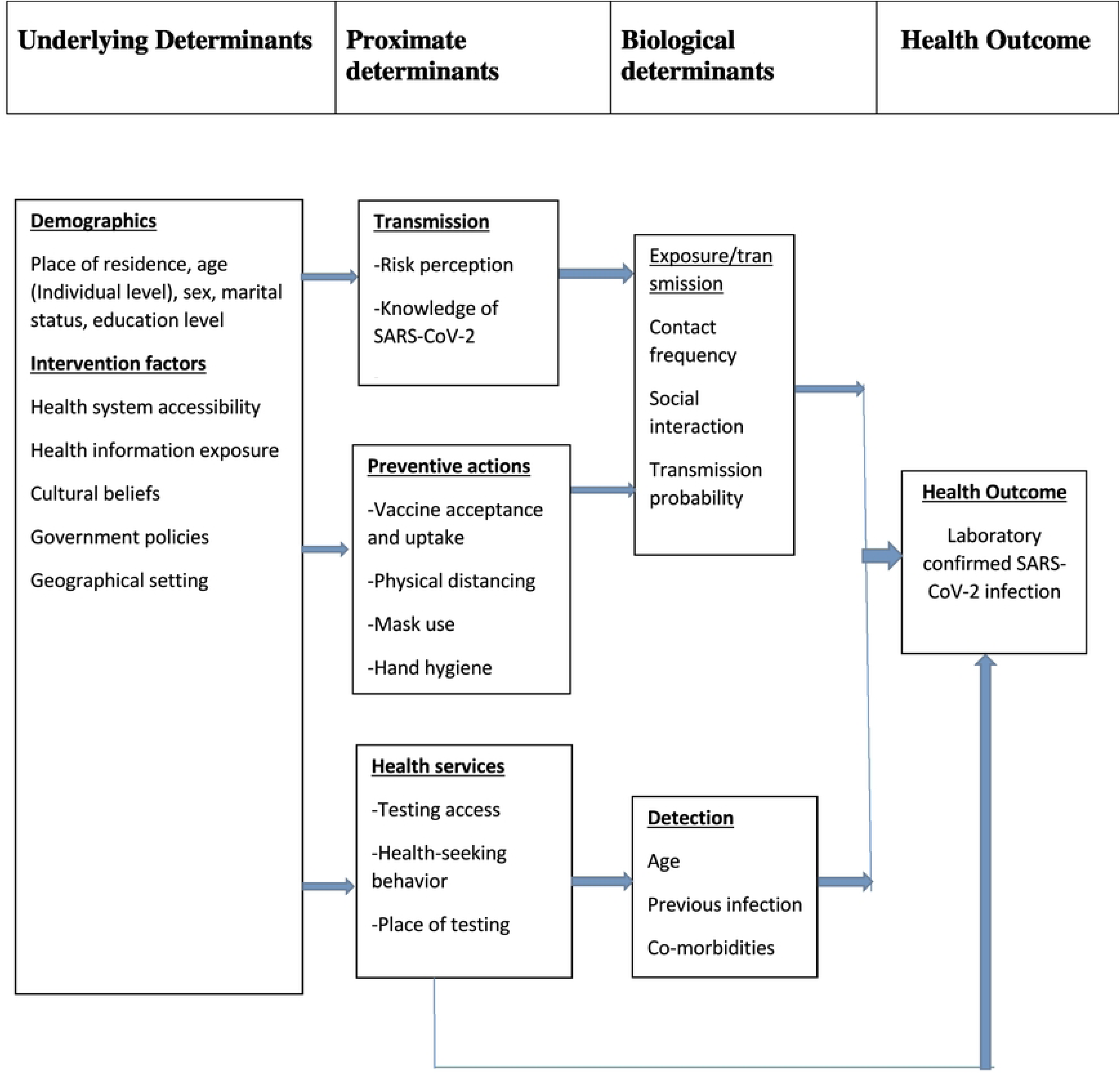
Conceptual framework for the underlying, proximate and biological determinants of SARS-CoV-2 infection

### Recruitment, sampling, sample size and data collection procedures

Recruitment of participants and data collection was conducted from 7^th^ October to 20^th^ December, 2024. Retrospective hospital-based surveillance data identified through record reviews covered period from 2019 to 2024, while primary data was collected between October and December, 2024. Community respondents were recruited directly from households.

Study employed different sampling strategies for Hospital-based surveillance data, community survey and qualitative data, Table 1. Retrospective surveillance data were obtained from identified Health facilities. These were selected purposively based on reported case burden. For each facility, patient records were reviewed and eligible cases selected systematically based on criteria. Due to use of retrospective surveillance data, sample size was determined by data availability. Multistage sampling technique was employed for the community survey where participants were drawn from the same residential areas. Firstly, all residential areas were selected within each district and eligible households randomly selected.

**Table 1:** Mixed-mode data collection and sampling techniques to recruit participants in Ndola, Lufwanyama, Kafue district of Zambia, October-December, 2024.

| Data Collection Mode | Target Number (n) | District/Areas | Sampling Techniques | Further Notes |
| --- | --- | --- | --- | --- |
| In-depth interviews (Face-to-face) | 30 | Ndola, Kafue and Lufwanyama residential areas (10 per site) | Purposive | Target population; Survivors of SARS-CoV-2 infection, Health care providers; Health administrators. |
| Hospital-based surveillance data | 395 | Ndola Central Hospital (L3); Lufwanyama District Hospital (L1); and Kafue General Hospital (L2) | Systematically | Reviewed SARS-CoV-2 patient files. Target of 132 per site. All active cases where included. |
| Community-based survey | 145 | Ndola, Kafue and Lufwanyama communities | Multistage | Community members |

Community survey sample size was estimated using Cochran’s formula assuming a 50% prevalence, 95% confidence level, design effect of 1.2 and 8% precision.

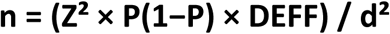

After adjusting for 20% non-response, the required sample size was 225 participants. However, due to logistical constraints, 145 participants were successfully enrolled.

For the qualitative component, 30 In-depth interviews with purposively selected insightful participants were conducted. Sample size was guided by the information-power approach and thematic saturation. Qualitative participants were not necessarily the same as those in the quantitative survey, but all met the eligibility criteria and were drawn from the same geographical settings. A summary of the sampling techniques and participant flow diagram is in Table 1 and Fig 2.

**Fig 2:**
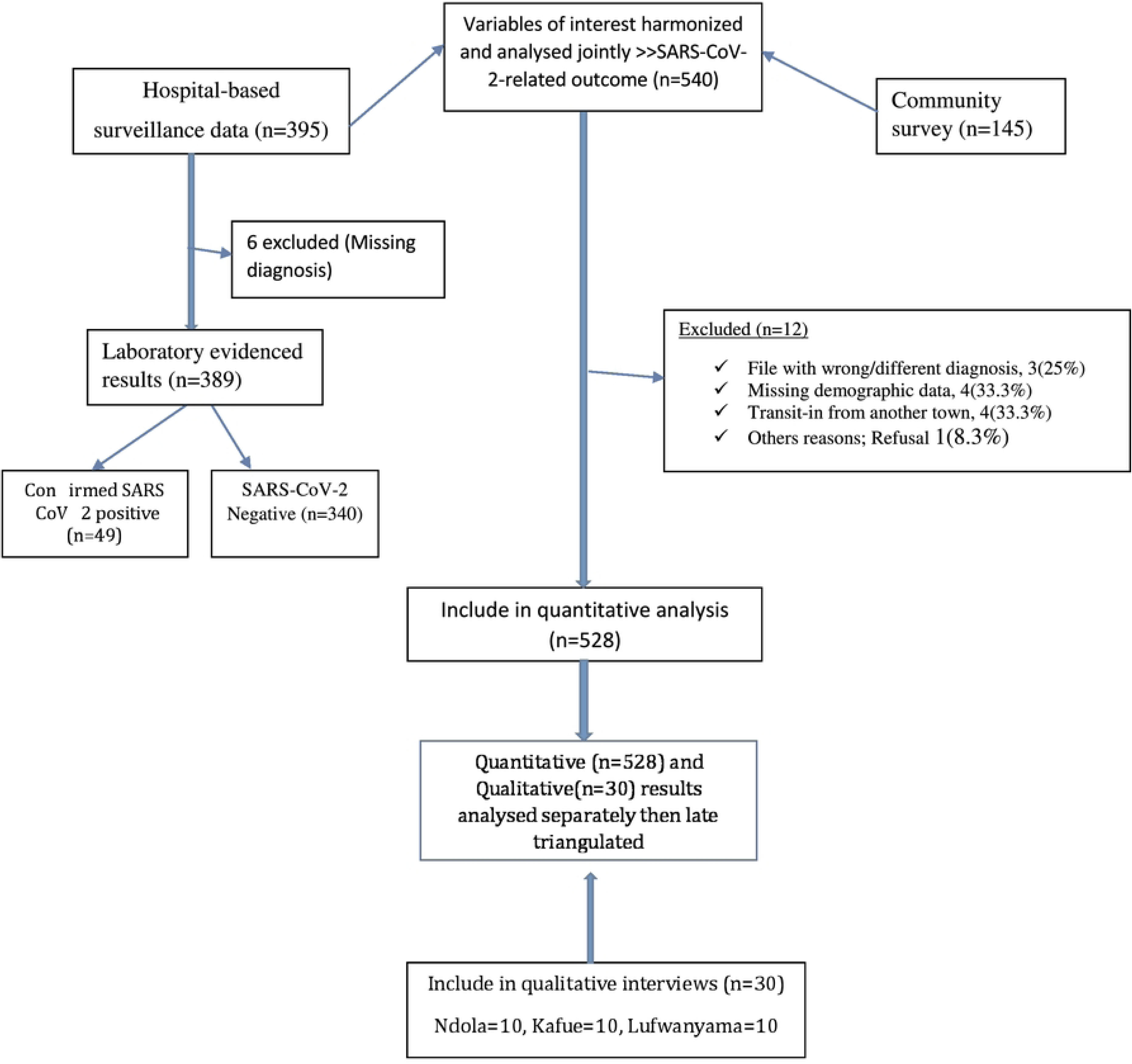
Diagram of participant recruitment, inclusion, sampling and analysis indicative per stage.

**Fig 3:**
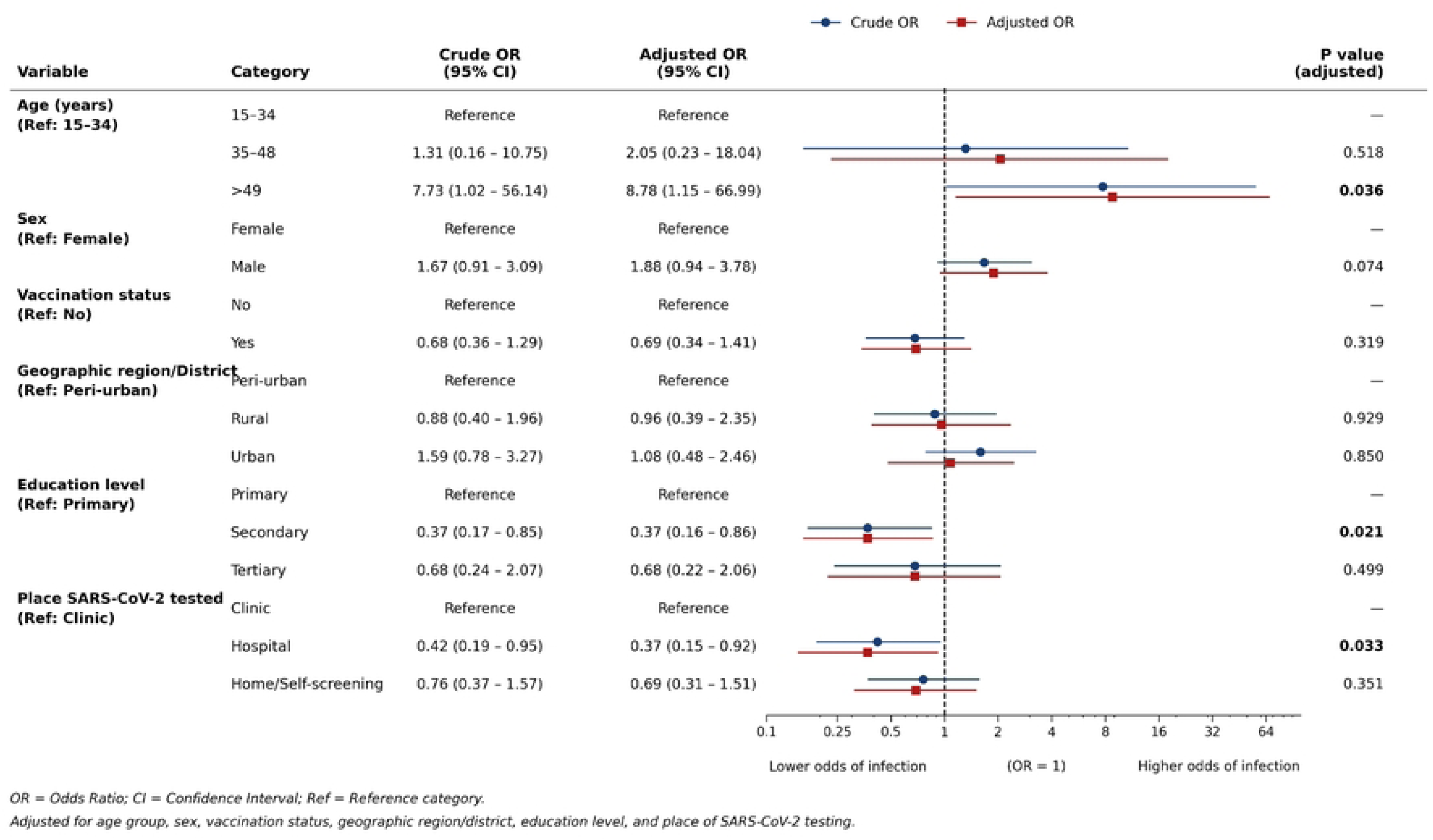
Crude and adjusted odds ratios for factors associated with SARS-CoV-2 infection (n=389)

**Fig 4:**
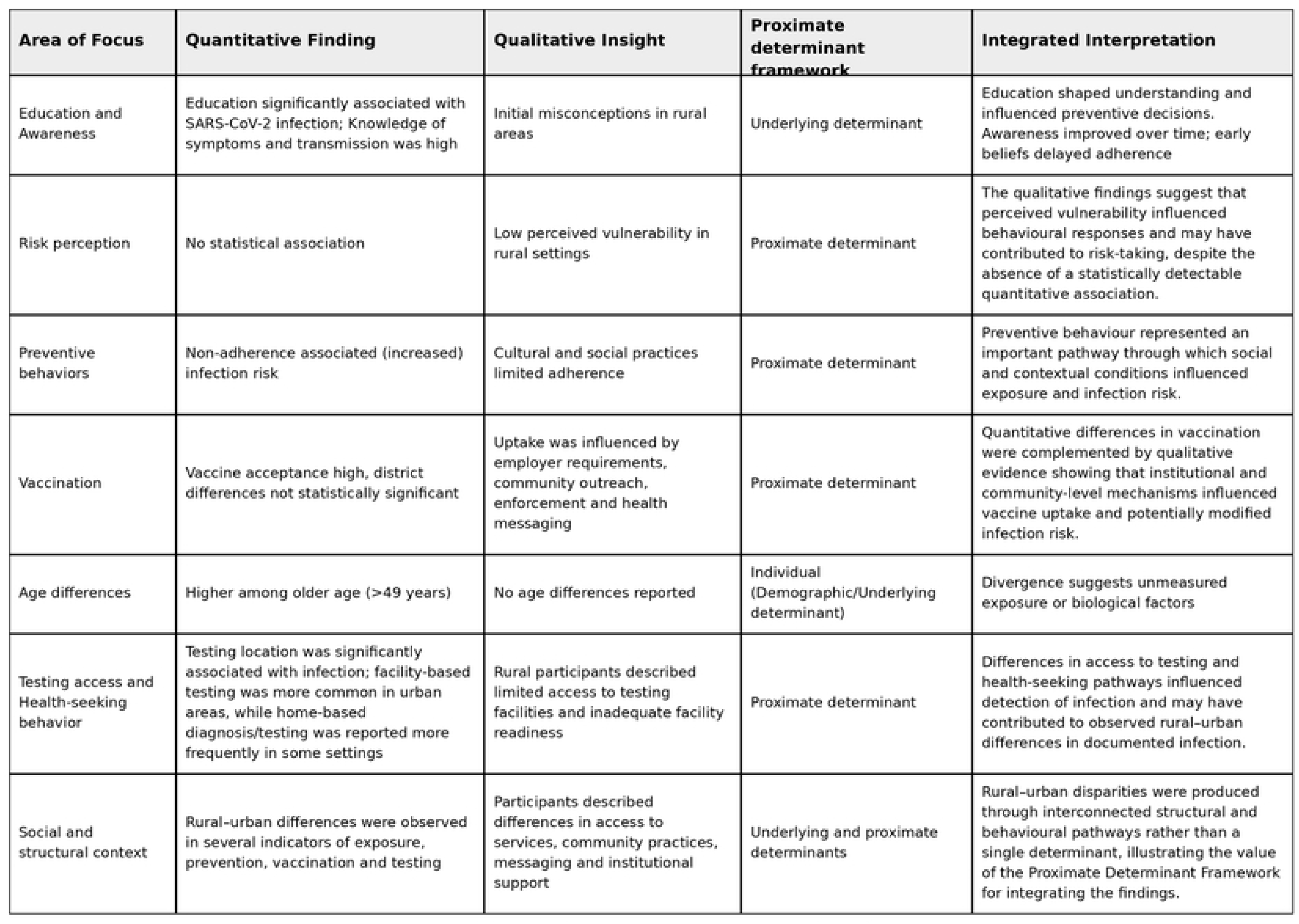
Triangulation of Quantitative and Qualitative Findings on Determinants of SARS-CoV-2 Infection Using the Proximate Determinant Framework

A structured questionnaire and a data extraction tool for hospital records were used to collect quantitative data. Variables of interest included; demographic characteristics, SARS-CoV-2 test results, clinical history, vaccination, experienced signs/symptoms. Qualitative data were collected using a semi-structured interview guide, with interviews conducted face-to-face. The questionnaire tool was developed by the research team after an extensive review of published literature, World Health Organization (WHO) COVID 19 surveillance tools, and study objectives cited as [Supplementary file 1]. The tools were pre-tested in 12 sampled participants in study setting, these individuals were excluded in the final analysis.

### Missing data, Statistical and thematic analysis

Quantitative data was analyzed using STATA version 17 (College Station, Texas, USA). Descriptive statistics summarized participants characteristics and SARS-CoV-2 case distribution across districts as frequency tables and cross tabulations. Bivariate analysis assessed associations between selected variables. Variables were selected a priori based on epidemiological relevance and the Proximate Determinant Framework, with consideration of confounding and collinearity. Such variables were included in multivariable logistic regression model to identify factors independently associated with primary outcome. Effect size reported using crude and adjusted odds ratios with 95% confidence intervals. The framework guided presentation of findings.

The two quantitative data sources (Community survey and Hospital-based surveillance) were harmonized by aligning variables of interest and analysed jointly to provide a comprehensive assessment of SARS-CoV-2-related outcome. Variables harmonized included; Age, district, sex, education level, place screened, marital status, vaccine acceptance and status. For logistic regression, only eligible hospital-based surveillance data were used.

Qualitative data was thematically analysed using framework described by Braun and Clarke (17). Interview transcripts were reviewed, coded manually and then organized into themes focused on perceptions and awareness of risk, prevention practices, and health-seeking behaviors. Quantitative and qualitative findings were then triangulated to enhance interpretation.

Prior to analysis, all datasets underwent data cleaning and validation. Records with incorrect diagnosis, substantial missing demographic information, or ineligible case classifications were excluded from analysis. Missing data were handled using complete-case analysis for variable-specific and multivariable analysis. Potential outliers were assessed through summary statistics, and graphical inspection, and no implausible values requiring removal were identified after verification against source records.

### Ethical considerations

Study obtained ethical approval from Chreso University Ethics Review Committee (Ref # 2254-09-2024) and National Health Research Authority (Ref # 2687/11/09/2024). Confidentiality and anonymity were maintained by removing personal identifies (Quantitative data) and restricting access to study data. After obtaining written informed consent a detailed personal structured questionnaire was carried out with all eligible members. For all face-to face interviews, written consent was obtained. For hospital reviews, verbal authorization was obtained from institution heads. The study obtained consent from parents for 5 minors aged between 15-17.

## RESULTS

### Participants Characteristics

540 participated in this mixed-method study conducted in three districts; Ndola (267), Lufwanyama (210) and Kafue (147). Urban participants (50.6%) constituted approximately half of the study population. The response rate was 78.9% with reasons for exclusion been; File with wrong/different diagnosis (25%); Missing demographic data (33.3%); Transit-in from another town (33.3%) and others i.e., Refusal (8.4%), Fig 2. More males 271 (51.3%) than females 257 (48.7%) participated with majority been single or divorced 269 (50.9%). The median age was relatively similar across districts, 31 (15–71). 4.3% (6/139) from community survey had no knowledge of SARS-CoV-2 transmission and prevention methods. Most participants trusted information about SARS-CoV-2 from health care providers or Health facilities (47.2%), radio or other social media (32.5%) and 20.3% from Ministry of Health website.

### SARS-CoV-2 infection and health services by geographical setting

Among eligible participants from hospital surveillance data, forty-nine (49/389), 12.6% had laboratory confirmed SARS-CoV-2 positive test. Of which, 57.1% (28/49) had a positive antibody/nasal swab result in their records while 42.9% (21/49) had a confirmed positive SARS test done. 42.9% (21/49) were active SARS-CoV-2 cases found during data collection; 20 cases in Ndola, 15 cases in Kafue and 14 cases in Lufwanyama (Table 2 and 3). Among confirmed active cases (n=21), 27.4% perceived no chance of infection, 18.7% very small chance and 53.9% as medium chance of contracting SARS-CoV-2 infection. SARS-CoV-2 case mortality was higher amongst the 40–49 years, majority who were in rural areas 2/14 (14.3%).

**Table 2:** Social-demographic characteristics, SARS-CoV-2 positive cases and testing services stratified by district, 2024 (n=528).

|  | Urban | Peri Urban | Rural |
| --- | --- | --- | --- |
| Characteristic | Ndola | Kafue | Lufwanyama |
|  | n=267 | n=147 | n=114 |
| Female, n (%) | 97(36.3) | 82(55.8) | 78(68.4) |
| Males, n (%) | 170(63.7) | 65(44.2) | 36(31.6) |
| Age (Years), mean (SD)* | 32(10.1) | 31(9.9) | 33(9.9) |
| Married/Cohabiting, n (%) | 122(45.7) | 47(31.9) | 90(78.9) |
| Vaccinated 2<sup>nd</sup> dose, n (%) * | 171(64.0) | 80(54.4) | 62(54.4) |
| Place screened, n (%) |  |  |  |
| Home-based/Self | 128(47.9) | 83(56.4) | 44(38.6) |
| Clinic | 103(38.5) | 46(31.3) | 30(26.3) |
| Hospital | 36(13.4) | 18(12.2) | 40(35.1) |
| SARS-CoV-2 tested positive | 20(7.5) | 15(10.2) | 14(12.3) |

**Note:** \* Differences in sample size based on complete available data for specific analysis. Detailed summary of SARS CoV-2 sero status in Table 3.

**Table 3:** SARS-CoV-2 infection positivity by District, 2024.

| District | Positive (n) | Tested (n) | Positivity (%) | 95% CI |
| --- | --- | --- | --- | --- |
| Ndola (Urban) | 20 | 203 | 9.9 | 0.06-0.15 |
| Kafue (Peri-urban) | 15 | 101 | 14.9 | 0.86-0.23 |
| Lufwanyama (Rural) | 14 | 85 | 16.5 | 0.09-0.26 |
| *Total SARS-CoV-2 cases, n=49. Positivity= number of positive SARS-CoV-2 tests/Total number tests performed, |  |  |  |  |

Participants reported different testing pathways during the pandemic. Overall, approximately half 255 (48.3%) reported self-screening while slightly more than half 273 (51.7%) utilized established formal health facilities. Significant geographic testing variation was observed in the districts. For instance, rural participants utilized more formal health facilities (61.4%) compared to peri-urban (43.5%) and urban participants (51.9%). Conversely, peri-urban participants (56.4%) had more home screening compared to rural (38.6%) and urban (47.9%) participants.

Vaccination represented one of the most important public health intervention programs during the pandemic. Analysis of vaccination records revealed substantial variation across districts. Urban participants 171/267 (64.0%) had the highest vaccination uptake compared to peri-urban (54.4%) or rural (54.4%), Table 2. Vaccine acceptance did not differ across districts. 244 (53.6%) agreed vaccines were safe and beneficial.

### Factors associated with laboratory-confirmed infections

#### Quantitative and Qualitative findings: SARS-CoV-2 infection and associated factors

Bivariate analysis suggests that age (>49 years), education and place SARS-CoV-2 screened were associated with SARS-CoV-2 infection status (Table 4). Participants aged 49 years and above had 8.8 times higher odds of infection compared to younger aged groups although the estimates were imprecise (aOR 8.78, 95%CI 1.15,66.99 P<0.036). No significant association between infection status and sex observed (P=0.126).

**Table 4:** Thematic analysis of risk perception, preventive behaviors and health-seeking behaviors of SARS-CoV-2 infection in selected districts of Zambia, 2024.

| Main Theme | Subtheme/Code | Code Description | Participant Quote | Interpretation |
| --- | --- | --- | --- | --- |
| Awareness and Perceived Risk | Knowledge of SARS-CoV-2 infection | Understanding of transmission, symptoms, and prevention | *“At first, people thought COVID was for those in towns or abroad, not for us in the village.” (Lufwanyama)* | Low early awareness affected adoption of preventive behaviors. |
|  | Perceived vulnerability | Beliefs about personal or family risk | *“I didn’t think I could get it because I am always at the farm and not in crowded places.” (Kafue participant)* | Risk perception shaped prevention practices. |
| Risk perception and Preventive Behaviors | Mask use and hygiene | Frequency and attitudes toward mask-wearing and handwashing | *“We wore masks when going to the clinic but not at home because it was uncomfortable.” (Ndola participant/caregiver)* | Preventive measures depended on context and enforcement. |
|  | Physical distancing | Practices in markets, homes, or gatherings | *“During funerals, we couldn’t keep distance; people said it’s disrespectful.” (Lufwanyama respondent)* | Cultural and social norms limited physical distancing. |
|  | Vaccination | Vaccine source | *"People were saying the vaccine causes infertility, so many refused."* | Hesitancy affect uptake |
|  |  | Public health requirement | *"My employer said we should get vaccinated if we wanted to continue working."* | Demand for normal-life enhanced uptake |
| Health-Seeking Behaviors | Health service access | Availability of clinics and response readiness | *“In our area, the clinic had no COVID kits; we were told to just isolate at home.” (Lufwanyama participant)* | Weak local health infrastructure readiness affected response. |
| Coping and Resilience | Adaptation and learning | Changes in practices over time | *“After seeing others fall sick, people started taking it seriously.” (Ndola youth)* | Experience increased knowledge and preventive action. |

Study observed no significant association between vaccination status and SARS-CoV-2 infection status (P≤0.319). Despite higher urban vaccine uptake, vaccination uptake remained insignificantly associated to SARS-CoV-2 infection status (cOR 1.59 95% CI 0.78 3.27, P<0.201). Study found a significant association between infection status and education level (P<0.021). Secondary education was associated with lower odds (aOR=0.37, P<0.021) of infection compared with primary education, whereas the association for tertiary education was not statistically significant (aOR=0.68, P<0.499).

Male participants had 1.7 times higher odds of SARS-CoV-2 positivity compared to females (cOR 1.88 95% CI 0.94,3.78), though statistically insignificant. Findings suggests a significant association between place of SARS-CoV-2 screening and primary outcome (aOR 0.37, 95%CI 0.15,0.92 P<0.033). Asymptomatic transmission was not analysed in our present study, though 7.9% tested positive asymptomatically; 2.8% (urban),5.1% (peri-urban) and none from rural. The common reported signs/symptoms among 21 active cases; Headache (32.4%), chills (21.2%), fever (23.9%) and cough (22.5%).

No significant difference in positivity of those tests conducted in urban and rural areas (χ²=3.4, P<0.132). Other demographic characteristics such as marital status and district (Location) remained insignificant. Given the small number of SARS-CoV-2 positive cases, some estimates showed wider confidence intervals, indicating limited precision. The final multivariable model “best fit model” had; age, education level, and place screened as significant factors associated with SARS-CoV-2 infection in this population. Multicollinearity among independent variables was assessed before modeling fitting using the Variance Inflation Factor (VIF), and model goodness-of-fit using Hosmer-lemeshow test to ensure adequacy of the final model.

In the qualitative findings, participants generally demonstrated awareness of SARS-CoV-2 infection and prevention measures. However, significant differences emerged in how risk was perceived across rural, peri-urban, and urban settings (Fig 4). Rural participants (n=8) consistently expressed perceived low risk to SARS-CoV-2 infection compared to urban participants. For instance, one rural participant expressed belief and said;

> *“Coronavirus is for town people; we live far, and it can’t reach here.”*

Participants described varying perceptions regarding personal susceptibility to SARS-CoV-2 infection. Several rural and peri-urban respondents initially believed they were unlikely to become infected. This was markedly among participants with lower education levels, who commonly perceived themselves as low risk. This was particularly so also among those residing or working in less crowded environment, such as farms. One participant stated;

> *” I didn’t think I could get it because I am always at the farm and not in crowded places.” (Kafue district)*

#### Quantitative and Qualitative findings: Vaccine acceptance and uptake

Vaccine acceptance did not differ significantly between the districts. 244 (53.6%) agreed that vaccines were good and safe, while 142 (31.2%) disagreed with the rest remaining neutral. Despite the high acceptance level, marked region variation in uptake was observed. Vaccine uptake was higher in urban 171/267 (64.0%) compared to rural areas 62/114 (54.4%). Majority vaccinated in urban populations were predominantly male and only 9/21 (42.9%) among active cases were vaccinated.

In bivariate analysis, there was significant association between vaccine acceptance and vaccine uptake (χ²=32.5, P<0.001). Urban participants had 1.2 times higher odds of being vaccinated compared to rural and peri-urban participants, though statistically insignificant (cOR=1.2,95%CI:0.73–2.07). Study observed no association between vaccination status and SARS CoV-2 infection status (aOR=0.69,95%CI:0.34–1.41).

Qualitative findings suggest that communities understood the importance of vaccination (n=26). Reasons for vaccine uptake differed across settings; in urban areas uptake was driven by employment requirements, travel needs, and greater exposure to messaging by Ministry of Health and other partners. One participant said;

> *“Here in Kafue, the government people would go around with mega phones to encourage people to go for vaccination, so people can go back to their normal businesses. I want for vaccination, because I wanted to go back to the market.”*

In rural areas on the other hand, uptake was mostly influenced by outreach campaigns, convenience, and community leader endorsement. In peri-urban settings, vaccine uptake had mixed influences which included workplace policies and community norms. Other prominent factors such as hesitancy also differed between rural and urban settings. In rural areas, fear of side effects and misinformation were the main drivers while distrust of government or pharmaceutical companies and sources of the vaccine were the main drivers in urban uptake.

#### Quantitative and Qualitative findings: Prevention behavior and adherence

Participants reported variation in preventive behaviors and practices during the pandemic. These behaviors included; mask wearing, hand washing, physical distancing and avoidance of crowded gatherings. There was no significant association between perceived self-risk and prevention practices (χ²=1.70, P<0.426). From community survey, 54 (43.6%) agreed adhering to preventive guidelines with 36 (29.0%) disagreeing and 34 (27.42%) remaining neutral. Among adherent group, only 26 (63.4%) were vaccinated. No significant association observed between vaccination status and adherence to prevention practices (χ²=1.27, P<0.530).

Qualitative findings varied substantially across settings and over time. Mask use was one of the most commonly reported preventive measure despite majority reporting that adherence was inconsistent. Physical distancing and hand washing were among widely promoted and most difficult preventive measures to monitor. Some facilities played an important role in reinforcing hygiene practices and social distancing as evidence by one urban participant who said;

> *“At the clinic they would not attend to you without washing your hands first and wearing a mask.”*

### Pragmatic triangulation with Proximate Determinant Framework synthesis: SARS-CoV-2 infection, vaccine and prevention-adherence practices

The integrated (18, 19) quantitative and qualitative findings demonstrate that SARS-CoV-2 infection was associated with a combination of demographic, geographical, behavioural and health-system factors rather than residence alone. Quantitative findings identified older age, education level and testing location as important factors associated with infection, while qualitative findings complemented and expanded these findings by illustrating how differences in risk perception, health literacy, healthcare access, testing behaviour, preventive practices and vaccination experiences varied across rural, peri-urban and urban communities. The integration of the two datasets demonstrated areas of convergence, complementarity and divergence, Fig 4. For example, the quantitative association between preventive behaviours and infection was supported by qualitative accounts of social and cultural constraints to adherence, whereas the absence of a significant quantitative association between risk perception and infection was expanded by qualitative evidence describing low perceived vulnerability in some rural communities.

Similarly, the quantitative association between testing location and infection was complemented by qualitative accounts of limited testing access and facility readiness. The Proximate Determinant Framework provided an analytical basis for integrating these findings by demonstrating how underlying determinants, such as education and geographical context, may operate through proximate determinants, including preventive behaviours, vaccination, risk perception and access to testing and healthcare, to shape SARS-CoV-2 infection patterns. Age represented an individual/demographic determinant associated with infection, although the qualitative findings did not provide a corresponding explanation for this association, indicating an area of divergence. Overall, the findings suggest that rural–urban differences in SARS-CoV-2 infection should be understood as the product of interconnected structural, behavioural and health-system pathways rather than geographical residence alone.

## DISCUSSION

The triangulated findings suggest that rural-urban differences in SARS-CoV-2 infection are better understood as differences in the social and health-system environments within which exposures and prevention occurs, rather than effect of geographical residence alone. This interpretation is consistent with evidence within sub-Saharan Africa that SARS-CoV-2 infection prevention has largely been shaped by interconnected factors (3, 5, 6, 20, 21); social economic, demographic, behavioral and health system factors. Studies across the region have demonstrated similarly that higher education attainment and knowledge do not necessarily translate to consistent behavioral practices (22, 23). Kaiser et al (2024) argue that personal choices and social expectations heavily dictated whether people followed safety steps. This indicates that willingness to act on health information is influence by broader context in which individuals live in. These findings support the framework by positioning education and geographical context which may operate through behavioral and health-system pathways associated with infection. Zhou et al (2021) further provided evidence on how social behaviors impact SARS-CoV-2 transmission (7)

The relationship (Fig 4) between knowledge, risk perception and behavioral is important. This is so because evidence from sub-Saharan and other parts of Africa has shown that although knowledge on SARS-Co-2 transmission and prevention are generally high (24, 25), perceptions of susceptibility and adherence to preventive measures vary considerably between populations. Rural populations often perceived themselves as low risk during outbreaks. The qualitative findings in this study help explain why knowledge alone maybe insufficient to prevent infection. Factors such as perceived vulnerability, social expectations and how feasible it is to maintain recommended behaviours likely influence how information was translated into action. This is substantial as it highlights that information disseminated may have limited impact where communities face social or practical constraints to adherence (26). Future outbreak communications should therefore combine accurate information with context-specific behaviour change strategies and community engagement.

Testing and healthcare access provided another additional explanation why the observed geographical disparities exist. Evidence from regional studies (4, 27, 28) have highlighted persistent inequalities in access to health-services and diagnostic capacity between urban-rural settings. Such inequalities have implications not only for access to care but also to visibility of disease within surveillance systems. Testing location was associated with laboratory-confirmed infection, potentially reflecting differences in health-seeking, referral and case ascertainment The qualitative evidence in this study suggests that geographical differences in documented infections may reflect differences in ascertainment and exposure. This is important because surveillance systems that largely depend on facility-based testing may systematically under-detect infections in communities with limited diagnostic capacity. Strengthening community-based surveillance, testing and approaches, decentralizing diagnostics capacity can be important to improve outbreak detection, reporting and reduce geographical surveillance biases.

The triangulation findings further highlight the importance of institutional and community influences on vaccination. Available evidence (29–31) indicates that vaccine acceptance and uptake are influenced not only by individual attitudes but also by trust, information, access, social influences and institutional factors. The qualitative evidence in this study seems to point to the importance of outreach, workplace requirements and public-health messaging as key drivers. This suggests that improving vaccine uptake during future outbreaks will require more than demonstrating vaccine safety and efficacy. It will require accessible delivery systems, trusted messengers and locally supported communications particularly in settings where health-services are limited.

An important contribution of this study is the integration of pragmatic evidence using the Proximate Determinant Framework. The framework has demonstrated that underlying determinants require biological and proximate-determinates to affect outcome. For instance, preventive behaviour represents an important pathway through which social and contextual conditions influenced exposure and infection risk. Secondly, mixed-methods analysis is crucial as it identifies areas where statistical and experiential evidence do not fully converge. The absence of a quantitative association between risk perception and infection, despite qualitative accounts of low perceived vulnerability, may indicate that perceived risk operates indirectly through behaviour or varies over time.

Likewise, the association between age and infection was not clearly explained by the qualitative data. Rather than treating these differences as weaknesses, they demonstrate that statistical measures may not capture the temporal, social and contextual dimensions through which determinants operate. This reinforces the value of mixed-methods approaches for infectious-disease research, particularly where behavioural and health-system processes are difficult to reduce to individual-level variables. Another strength was integrating hospital surveillance data, community survey findings and qualitative interviews. Study included rural, peri-urban and urban populations providing important comparative evidence from different epidemiological settings.

Application of the Proximate Determinant Framework strengthened interpretation by linking demographic, behavioural and structural determinants to infection risk. However, several limitations should be acknowledged. The relatively small number of laboratory-confirmed cases reduced statistical precision for some regression estimates, resulting in wide confidence intervals. The cross-sectional nature of the community survey limits causal inference, while retrospective hospital surveillance data depended on completeness of routinely collected records. The study was also conducted during the post-pandemic period when infection prevalence had declined, limiting opportunities for detailed spatial and temporal analyses.

Taken together, the findings support a shift from viewing rural–urban disparities as a simple geographical phenomenon towards understanding them as a manifestation of unequal distribution of information, health-system capacity, preventive opportunities and diagnostic access. This interpretation has practical implications for Zambia’s preparedness for future similar epidemics. A geographically uniform intervention is unlikely to produce equitable outcomes where communities differ substantially in their ability to access information, testing, vaccination and healthcare. Public-health preparedness should therefore combine context-specific risk communication, community-based prevention, equitable vaccination delivery, decentralized diagnostic services and strengthened rural surveillance. The Proximate Determinant Framework provided a useful basis for understanding how these upstream and proximate factors interact to shape infection risk and detection. Furthermore, integrating socio-economic and behavioral determinants into outbreak response planning can support more effective and sustainable disease prevention strategies.

## CONCLUSION

Study has demonstrated that Laboratory-confirmed SARS-CoV-2 infection continued to be detected in the study settings during the post-peak pandemic period and was different across rural, peri-urban, and urban settings. Infection was independently associated with age, education and testing location, while vaccination uptake differed geographically but was not associated with infection. Qualitative findings highlighted behavioral-contextual and health-system factors affecting transmission. Despite population being knowledgeable and aware about SARS-CoV-2 infection, gaps in prevention adherence, and access to testing services particularly in rural areas remain a challenge. Strengthening routine surveillance, access to testing services and vaccination, and implementing contextual-specific prevention strategies in high-risk populations are key to enhance preparedness and emergency responses to SARS-CoV-2 and future similar respiratory outbreaks.

## Data Availability

Data availability requirements Our study involved human participants and includes sensitive information related to SARS-CoV-2 symptoms, testing experiences, demographic, and health service use across three districts. De-identified quantitative data set labelled, ‘S2 Quantitative data set” has been submitted as part of supplementary files. The identifiable qualitative data set (in-depth interviews) which are raw audio recordings contain potential identifying and sensitive information such as age, location, sex, and occupation. In accordance with our approved ethical protocol, National Health Research Authority (NHRA) and Chreso University Ethics Review Committee (CUREC) do not permit public posting of raw qualitative audio recordings, as these contain voices and contextual information that pose a risk of participant re-identification. To comply with these restrictions, we have provided fully de-identified qualitative transcripts/excerpts labelled, S3 file de-identified qualitative excerpts as part of the supplementary files. These excerpts represent the minimal dataset required to understand and verify the qualitative findings without compromising participant confidentiality. a) Ethical Restrictions The raw qualitative audio files contain potentially identifying and sensitive health information, including unique verbal characteristics, local details, and personal experiences that cannot be fully anonymized. For this reason, both NHRA and the CUREC require that such data be shared only upon reasonable request and not uploaded to public repositories. Independent, non-author bodies responsible for reviewing data access requests are: Chreso University Ethics Review Committee (CUREC) Phone: +260 211355590 National Health Research Authority (NHRA) Website: https://nhra.org.zm Researchers requesting access must provide: 1. A formal request letter 2. Proof of institutional ethics approval 3. A data use agreement ensuring confidentiality 4. A brief description of intended use and analyses b) Data Ownership All data were collected by the authors but remain under the regulatory oversight of NHRA and CUREC. External researchers may request access through the procedures above. The authors did not have any special access privileges beyond standard approval processes. c) Updated Data Availability Statement The minimal dataset necessary to understand and replicate the qualitative findings has been provided in the form of de-identified transcripts/excerpts. Raw audio recordings contain identifiable human participant information and cannot be publicly shared due to ethical restrictions imposed by the National Health Research Authority and CUREC. De-identified data will be made available upon reasonable request to the CUREC or to NHRA for researchers who meet the criteria for access to confidential data.

## Supporting information

S1 file Data availability requirements

S2 File Quantitative data set

S3 file date file-De-identified qualitative transcripts/excerpts

S4 file Data collection tools

S5 file National health Research Authority (NHRA) approval letter

S6 file Ethics review committee approval

S7 Fig 1: Conceptual framework for the underlying, proximate and biological determinants of SARS-CoV-2 infection

S8 Fig 2: Diagram of participant recruitment, inclusion, sampling and analysis indicative per stage.

S9 Fig 3: Crude and adjusted odds ratios for factors associated with SARS-CoV-2 infection (n=389)

S10 Fig 4: Triangulation of Quantitative and Qualitative Findings on Determinants of SARS-CoV-2 Infection Using the Proximate Determinant Framework

## Notes

### Competing Interest Statement

The authors have declared no competing interest.

### Author Declarations

Chreso University Ethics Review Committee and Zambia National Health Research Authority

